# Sub-spreading events limit the reliable elimination of heterogeneous epidemics

**DOI:** 10.1101/2021.03.13.21253477

**Authors:** Kris V Parag

## Abstract

We show that sub-spreading events i.e., transmission events in which an infection propagates to few or no individuals, can be surprisingly important for defining the lifetime of an infectious disease epidemic and hence its waiting time to elimination or fade-out, measured from the time-point of its last observed case. While limiting super-spreading promotes more effective control when cases are growing, we find that when incidence is waning, curbing sub-spreading is more important for achieving reliable elimination of the epidemic. Controlling super-spreading in this low-transmissibility phase offers diminishing returns over non-selective, population-wide measures. By restricting sub-spreading we efficiently dampen remaining variations among the reproduction numbers of infectious events, which minimises the risk of premature and late end-of-epidemic declarations. Because case-ascertainment or reporting rates can be modelled in exactly the same way as control policies, we concurrently show that the under-reporting of sub-spreading events during waning phases will engender overconfident assessments of epidemic elimination. While controlling sub-spreading may not be easily realised, the likely neglecting of these events by surveillance systems could result in unexpectedly risky end-of-epidemic declarations. Super-spreading controls the size of the epidemic peak but sub-spreading mediates the variability of its tail.

## Background

Emerging infectious diseases are a major and recurring threat to both global health and economies. The ongoing COVID-19 pandemic has exemplified this, highlighting a need for improved understanding on how interventions might be applied and relaxed to jointly minimise public health risks and socio-economic costs. Key parameters, which characterise the impact of interventions on the transmission dynamics of an infectious disease are the time-varying event reproduction number, denoted *R*_*s*_ at time *s*, with mean *µ*_*s*_, [1] and the dispersion level, *k*, of the offspring distribution of the epidemic [2]. The first describes transmission potential by measuring the number of secondary infections per primary case at *s*. The second defines transmission heterogeneity i.e., it captures the variation of possible *R*_*s*_ about mean *µ*_*s*_. The values of these parameters often inform intervention policy and much debate remains on their implications for both epidemic control and elimination [3], [4], [5].

A *µ*_*s*_ *>* 1 forewarns of rising incidence (new cases), necessitating the application of controls, while *µ*_*s*_ *<* 1 signifies that the epidemic is being controlled, potentially allowing the relaxation of some interventions [1]. Moreover, the risk and effectiveness of policies are modulated by *k*. If *k* ≫ 1 then transmission is homogeneous (*µ*_*s*_ is representative of the realised *R*_*s*_) and we can apply simple, population-wide controls. Alternatively, if *k <* 1, then heterogeneity is large (the epidemic offspring distribution is overdispersed). This means that infrequent super-spreading events, in which each primary case causes ≫ *µ*_*s*_ secondary ones, disproportionately drive overall transmission [2]. The majority of other transmission events, which we refer to as sub-spreading events, lead to fewer than *µ*_*s*_ or even 0 secondary cases. Heterogeneous epidemics may require more targeted, event-specific policy as population-wide measures can often be ineffectual [2], [6].

Many works have examined how *k* and *µ*_*s*_ interact to regulate the effectiveness of control measures applied to growing epidemics. When transmission is heterogeneous, the consensus is that interventions aimed at mitigating super-spreading events (e.g., by limiting large gatherings) can minimise both peak epidemic size and resource-usage [2], [7], [8]. While these insights inform on how to efficiently impose control measures, the converse problem of when to safely release interventions and de-escalate surveillance, for waning epidemics, has been understudied [9], [10]. The importance of this problem will only elevate as countries debate the value of elimination-based strategies for handling the ongoing COVID-19 pandemic [3], [11]. In this paper we investigate this relaxation problem in the context of achieving reliable and minimum-risk end-of-epidemic declarations.

The timing of an end-of-epidemic declaration is strategically crucial as it triggers the removal of restrictions or control measures, the re-allocation of resources and the resumption of trade, travel and other economic activities. However, getting this timing right is both non-trivial and consequential. Early declarations (and hence intervention relaxation) can elevate the risk of resurgence or second waves, whereas late ones can cause needless cost. Much is unknown about how the transmission dynamics of an infectious disease dictate the tail of an epidemic. Recent studies suggest that this knowledge-gap may have contributed to early declarations for Ebola virus disease (EVD) in West Africa [12] and late ones for MERS-CoV in South Korea [13].

Prime among these unknowns is the influence of heterogeneity and its associated targeted interventions. While some studies have started analysing the interacting roles of *µ*_*s*_ and *k* to decipher this influence [12], [14], a general framework for rigorously testing hypotheses about how heterogeneity and control choices impact end-of-epidemic declarations is lacking. We extend theory from [2], [15] to develop such a framework, which allows us to generate novel and principled insight into how *µ*_*s*_ and *k* (and hence possible *R*_*s*_ values) mediate the tail of an epidemic. We take a bottom-up approach, tracing how heterogeneity in transmission, defined by variations in *R*_*s*_ due to *k*, causes fluctuations in incidence, which then manifest as noise in the probability of epidemic fade-out or elimination.

As this probability defines our confidence in any end-of-epidemic declaration [15], we obtain a measure of the risk of that declaration. This framework leads to several new results. First, we observe that while decreasing *k* (increasing heterogeneity) reduces the mean waiting time to elimination, measured from the last observed reported case (a standard reference point [16]), it also increases the maximum waiting time. This contextualises the common assertion that heterogeneity increases the likelihood of epidemic extinction [17], showing that while this may happen on average, the increased uncertainty it brings actually hampers safe end-of-epidemic declarations.

Second, we find that sub-spreading, surprisingly, plays a larger part than super-spreading in setting the uncertainty around the probability of elimination and hence end-of-epidemic declarations. We confirm this by testing three control strategies, which target (i) all transmission events, (ii) super-spreading and (iii) sub-spreading, for a fixed level of control effort *ρ* (the mean reproduction number is then *ρµ*_*s*_). Strikingly, we see that waning epidemics, which feature *ρµ*_*s*_ *<* 1, are most reliably eliminated by curbing sub-spreading events. Moreover, we find the benefits of super-spreading control relative to the simpler approach of (i) largely disappear in this epidemic phase.

Because *ρ* can equally model the mean case reporting or ascertainment rate [18], results from (i)-(iii) concurrently describe the impact of uniform, size-inverse (super-spreading events are under-ascertained) and size-biased or preferential reporting (sub-spreading is under-ascertained), respectively. Consequently, we uncover that the under-reporting of sub-spreading events, which is likely, can engender overconfident and risky end-of-epidemic declarations. These issues are in addition to known biases that under-reporting cause for epidemic elimination [15]. Robustly assessing the endpoint of an outbreak may therefore require either targeting interventions at sub-spreading events (if possible) or increasing contact tracing surveillance to minimise the under-reporting of those events.

One defining characteristic of heterogeneous transmission is an excess of sub-spreading events (zero-inflation) [2]. However, these events have received little attention in the literature. Only recently has the importance of sub-spreading been recognised, in related network epidemic models [19]. Our results draw attention to the unexpected influence of sub-spreading, clarifying its impact on epidemic elimination and adding context and detail to the subtleties of controlling and monitoring epidemics with overdispersed transmission. Hopefully, our framework will help inform guidelines for achieving safe and efficient end-of-epidemic declarations and intervention de-escalations in the face of heterogeneity and improve understanding of the complex dynamics driving real epidemics.

## Methods

### Renewal models with heterogeneous transmission

We consider an outbreak observed daily over the time period (in days) [1, *t*] with the number of newly infected cases on day *s* ≤ *t* as *I*_*s*_. The incidence curve of this epidemic is denoted 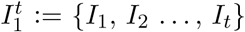. We model the time-varying transmission of a communicable disease within a population using a renewal process [20] [21]. This process describes how an infection spreads from a primary case to secondary ones at time *s* using two key variables: the effective reproduction number, *R*_*s*_, and the generation time distribution, with probability *w*_*u*_ at time *u*. Here *R*_*s*_ is the number of secondary cases at time *s* every effective case at *s* − 1 infects, while *w*_*u*_ gives the probability that the average time for a primary to secondary transmission is *u* days [20].

Under this renewal process framework, the observed incident cases at time *s* depend on *R*_*s*_ and past cases (over period [1, *s* − 1]) according to the Poisson (Pois) relation on the left side of Eq. (1).

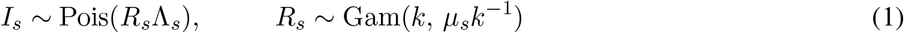

Here 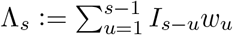, which depends on 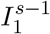, is known as the total infectiousness. It characterises how many past effective cases contribute to the next observed case-count at *s*. The generation time distribution (which we assume to be equal to the serial interval distribution) is central to defining the impact of each past case [20].

If we describe an epidemic as consisting of a sequence of spreading or transmission events, with the reproduction number of the spreading event at time *s* as *R*_*s*_ then standard renewal models assume a fixed *R*_*s*_ [21], [22], [23]. This formulation, while useful, does not account for possible heterogeneities in transmission, which are known features of many respiratory diseases such as the SARS and MERS coronaviruses [2]. If we define a distribution over *R*_*s*_ with mean 𝔼[*R*_*s*_] = *µ*_*s*_ then these models set 𝕡(*R*_*s*_ = *µ*_*s*_) = 1. Heterogeneous transmission is a mean preserving spread of this condition i.e., events with fixed mean *µ*_*s*_ can have different *R*_*s*_ values.

We define super-spreading events as those driven by *R*_*s*_ significantly larger than *µ*_*s*_. This characterisation differs slightly from the standard in [2], which directly uses numbers of secondary cases. Since the renewal model has long-term memory (i.e., factors in the age of infections via Λ_*s*_) using *I*_*s*_ would not be as appropriate here. However, because *I*_*s*_ behaves like a noisy, scaled version of *R*_*s*_ (by the properties of Poisson mixtures [24]), these two definitions are largely consistent. In this work we consider the end of the epidemic, which follows the waning phase of the epidemic. This contrasts the development in [2], which focusses on the growth phase. We define sub-spreading events as having *R*_*s*_ notably smaller than *µ*_*s*_. This type of event has received appreciably less attention (than super-spreading ones) and is our main topic of study.

If we make the usual assumption that *R*_*s*_ is gamma (Gam) distributed with shape *k* and scale *µ*_*s*_*k*^*−*1^ i.e., the right side of Eq. (1), then we obtain the negative binomial (NB) relation 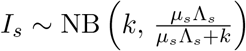 This is the most common method for incorporating heterogeneity within renewal model frameworks [12], [14], [25]. As *k* gets smaller the likelihood of both super-and sub-spreading events increases [2], [24]. Special cases are at *k* → ∞, *k* = 1 and *k* → 0 for which *I*_*s*_ has a Poisson, geometric and logarithmic distribution respectively with mean *µ*_*s*_Λ_*s*_ [18]. Many of the infectious diseases that feature significant heterogeneity have been found to exhibit *k <* 1 [2], [26]. This NB model can also be used to describe reporting noise and other types of heterogeneity.

### Variation in reproduction numbers and incidence

We explicitly characterise how heterogeneity in transmission can control the incidence of an epidemic and then assess the implications of this observation. Consider any arbitrary distribution over the effective reproduction number at time *s, R*_*s*_, with mean *µ*_*s*_. Using the law of total expectation we can write 𝔼_*I*_[*I*_*s*_] = 𝔼_*R*_𝔼_*I*_[*I*_*s*_ | *R*_*s*_] with the subscripts clarifying (when needed) about which variable we are taking expectations. Using the renewal model (left expression in Eq. (1)), we obtain the straightforward but general Eq. (2) below.

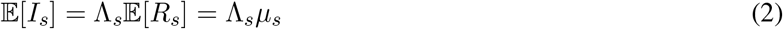

Eq. (2) shows that for any renewal model there is a direct relationship between mean incidence and *µ*_*s*_.

Similarly, we apply the law of total variance to get: 𝕧[*I*_*s*_] = 𝔼_*R*_[𝕧_*I*_[*I*_*s*_ | *R*_*s*_]] + 𝕧_*R*_[𝔼_*I*_[*I*_*s*_ | *R*_*s*_]] [24] (with 𝕧[.] indicating variance). Expanding this for renewal models we derive Eq. (3), which holds for any *R*_*s*_ distribution.

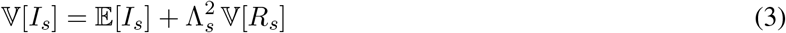

If we substitute the statistics from the standard gamma *R*_*s*_ distribution (right expression in Eq. (1)) we recover the usual variance of the NB incidence distribution i.e., 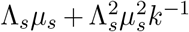 [12]. The key takeaway from Eq. (3) is that, given the past (summarised by Λ_*s*_), the variance of the reproduction numbers linearly controls that of the incidence at any time. Eq. (2) and Eq. (3) explain why we can generally map dynamics of *R*_*s*_ onto *I*_*s*_.

As a result, we can decipher the influence of super-and sub-spreading events (which are respectively linked to the tail and head i.e the right and left portions of the *R*_*s*_ distribution) by investigating the level of heterogeneity or variation among reproduction numbers. This is useful because it is hard to reliably estimate secondary cases caused by past primary cases during the waning stages of the epidemic [13].

The impact of control measures [2] or under-reporting [18] (which are often mathematical analogues) can be measured by their signature on the variance-to-mean ratio (VM) of the incidence: 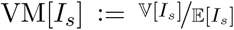 Using Eq. (2) and Eq. (3) we obtain Eq. (4) connecting the VM ratios of *R*_*s*_ to *I*_*s*_.

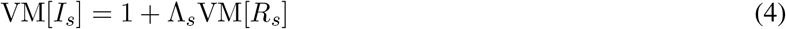

Thus, once again the statistics of *R*_*s*_ strongly modulate those of *I*_*s*_ at any time, for a given Λ_*s*_. While this result is simple, its ramifications, which are meaningful, have not been explored.

Understanding how properties of the *R*_*s*_ distribution map onto the related *I*_*s*_ one, which is a mixed Poisson distribution, provides insights into other epidemic properties. From mixed Poisson theory [24] we deduce that (a) if *R*_*s*_ is unimodal and continuous then *I*_*s*_ is also unimodal, (b) the shape of *I*_*s*_ will be similar to that of the mixing distribution describing *R*_*s*_ (e.g., when *R*_*s*_ is exponential, *I*_*s*_ becomes geometrically distributed) and (c) every mixed Poisson distribution corresponds to a unique mixing distribution i.e., multiple *R*_*s*_ distributions cannot map to the same *I*_*s*_ distribution [24]. Properties (a)–(c) establish that much about super-and sub-spreading events can be learned from *R*_*s*_. We will exploit these relationships to better understand epidemic elimination and heterogeneity.

### Elimination probabilities and epidemic lifetimes

We define an epidemic to be eliminated (or extinct) [5] at some time *s* if no future infected (local) cases are observed i.e., *I*_*s*+1_ = *I*_*s*+2_ = … = *I*_*∞*_ = 0 [5], [15]. If we initially assume that all future mean reproduction numbers, 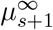, are known, then we can construct the probability of elimination given some sample 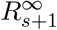 from 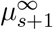 (see Eq. (1)) as 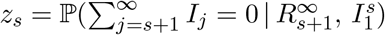. As we condition on the sample 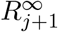 and because incidence is non-negative and *I*_*j*_ does not depend on 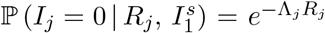, we can decompose *z*_*s*_ to get Eq. (5).

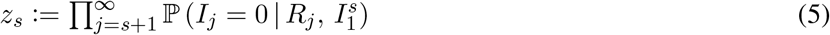

From Eq. (1), 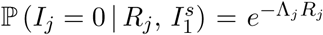 (also see [3]) and we obtain Eq. (6), with future Λ_*j*_ computed by incorporating the 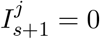 terms from above as pseudo-data (see [15], [27]).

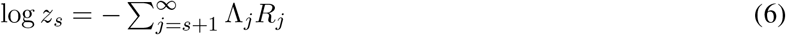

Hence, the elimination probability has an uncomplicated log-linear dependence on the sample sequence of reproduction numbers and accordingly depends on all future spreading events. This type of relationship is maintained even if we generalise Eq. (1) to include case observation noise (e.g due to importation or delays) [22], [28]. Methods for inferring *z*_*s*_ given these noise sources are currently being developed in [10], [15].

While Eq. (5) can be computed at any time, commonly elimination is only considered when zero-case days are observed in sequence [13], [16]. Keeping to convention, we will often present results in terms of Δ*s*, which is the time relative to that at which incidence was last non-zero, *t*_0_. We compute the relative time at which the epidemic is eliminated with *α*% confidence, given the transmission event sample 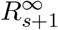, as *t*_*α*_ in Eq. (7).

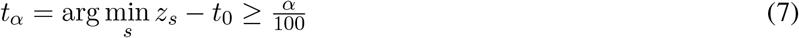

Eq. (7) also gives the time that an epidemic, composed of spreading events 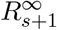, can be declared over with at least *α*% confidence [15], [29]. This confidence reflects the remaining variability among possible epidemic trajectories despite conditioning on the fixed future 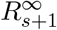 sample and past observed incidence 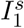.

If we think of the epidemic as a process that generates infections then its survival function, which captures the probability of the epidemic propagating at least 1 future case after *s*, is precisely 1 − *z*_*s*_. The mean lifetime of the epidemic then follows from survival theory (where it is called the mean time to failure) as 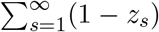 [30]. Given 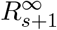, we can compute this lifetime exactly, as 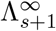 only depends on future incidence values, which are all 0 for this condition [15]. Hence *z*_*s*_ has an influential role in determining the risk of epidemic survival. As end-of-epidemic declarations times depend on *z*_*s*_, this risk has important public health and economic implications. However, we cannot know what sample from 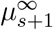 will be realised, and must account for this uncertainty, which derives from the heterogeneity in transmission and hence depends on the levels of super- and sub-spreading. Since, practically, an authority would want a single time for declaring an epidemic to be eliminated or over [16], we need some way of summarising the range of *z*_*s*_ curves and hence *t*_*α*_ values that could result. We propose two such statistics: the mean 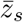 and the minimum *z*_*s*,min_. The resulting declaration times are then 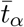 and *t*_*α*,max_. The mean elimination probability was used in [15], while the worst case (minimum) was proposed in [29].

We derive 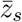 exactly from Eq. (6) as 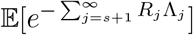. This yields Eq. (8) with 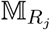 as the moment generating function according to the distribution of *R*_*j*_, which we evaluate at Λ_*j*_ [31].

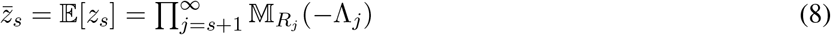

This formulation has immediate consequences [20]. First, 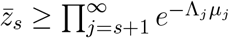. Ignoring heterogeneity will therefore on average result in our underestimating the elimination probability and so overestimating when the outbreak is over with *α*% confidence. This extends earlier works, which used simpler branching process epidemic models to link heterogeneity and extinction [2], to more realistic renewal epidemic descriptions [7].

Second, because 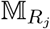 has a unique correspondence to the distribution over *R*_*j*_ [31], sub- and super-spreading events have direct roles in shaping the mean elimination probability. For example, given some threshold *c*, Chernoff’s bound stipulates that 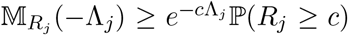, with 𝕡(*R*_*j*_ ≥ *c*) specifying our spreading event likelihoods. Last, under the gamma distribution in Eq. (1) we can explicitly compute Eq. (8) as Eq. (9) [31].

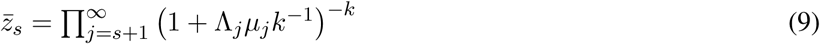

Eq. (9) provides intuition into how the offspring dispersion parameter *k* controls the mean elimination probability and hence the epidemic lifetime. Applying the product rule, we can show that 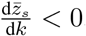, proving that as heterogeneity rises i.e., *k* gets smaller, 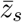 (and hence 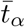) always increases (respectively decreases).

While the worst case elimination probability, *z*_*s*,min_, does not admit such analytic development and so is computed via simulation, we gain some insight about its behaviour from the variance among the *z*_*s*_ curves: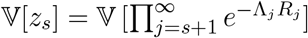. Each term in this product is independent so we decompose this to 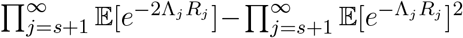 to get Eq. (10) with 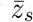 taken from Eq. (8).

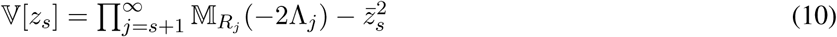

We obtain an explicit form for Eq. (10) by substituting the moment generating functions for gamma distributions as in Eq. (9). It follows that lim_*k→∞*_ 𝕧[*z*_*s*_] = 0. We generally find that as *k* falls, variation among *z*_*s*_ curves and hence possible *t*_*α*_ increases (see Results). The gap between *t*_*α*,max_ and 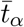 widens with heterogeneity.

Understanding how variation in *R*_*s*_ maps to uncertainty in elimination times is the main focus of this work. Specifically, we examine how changes to this variation, due to different control or case ascertainment strategies, express themselves in our ability to reliably adjudge the endpoint of the epidemic. In the Results we exploit the mathematical framework developed here to investigate how 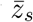 and *z*_*s*,min_, which are two measures of the aggregate risk of an epidemic, depend on super- and sub-spreading events. Based on these, we attempt to derive schemes for reliably eliminating an epidemic i.e., reducing its lifetime with minimum risk. This moment generating function approach to epidemic elimination or fade-out is novel, as far as we are aware.

## Results

### Reducing variation among reproduction numbers

In Eq. (4) we showed how VM ratios of the event reproduction numbers, *R*_*s*_, directly control those of the incidence values, *I*_*s*_. We first verify this relationship on numerous epidemics simulated according to the heterogeneous renewal model of Eq. (1). We consider epidemics characterised by an initial exponential growth followed by drastic control (e.g., a lockdown measure) and compute the VM ratios of both *I*_*s*_ and *R*_*s*_ for all times *s* of the epidemics. Fig. 1 shows that as heterogeneity increases (i.e., *k* falls) the VM of *I*_*s*_ and *R*_*s*_ both rise, despite different mean values across the possible epidemics as well as varying total infectiousness Λ_*s*_.

**Fig. 1:**
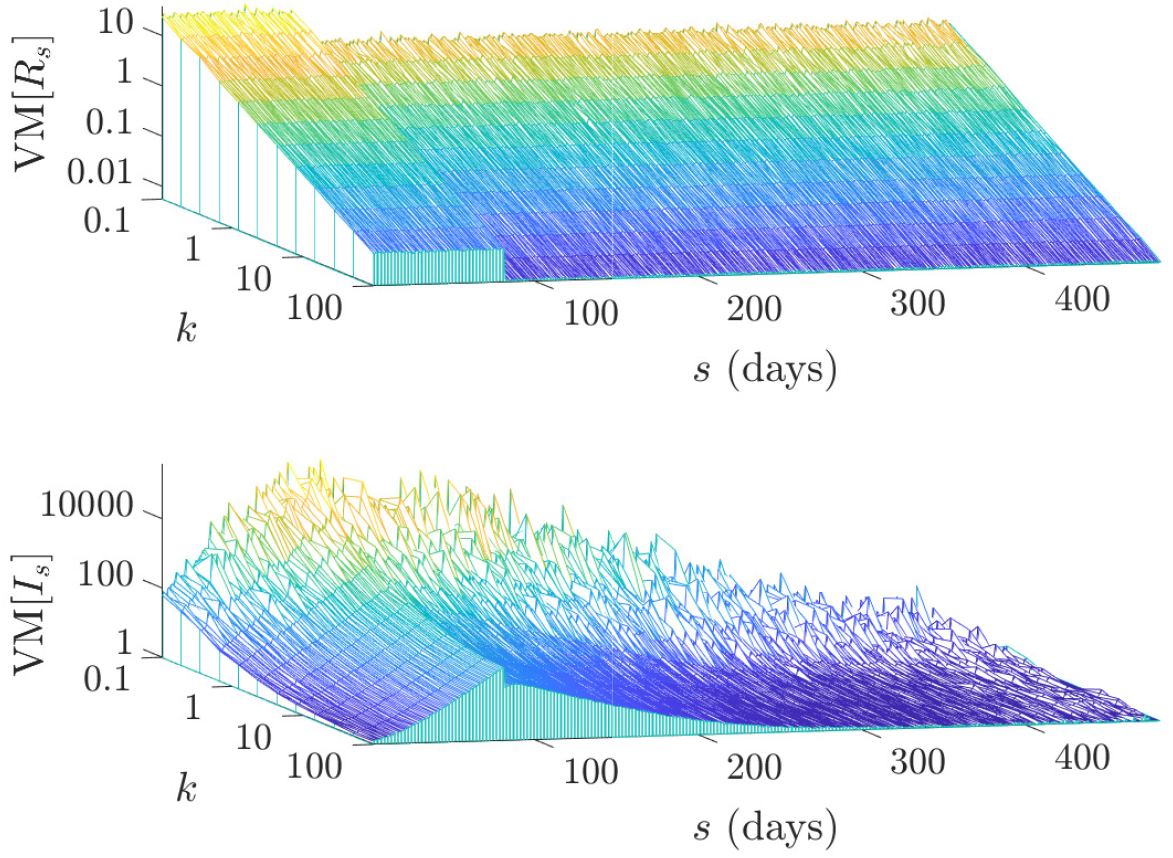
Variance-to-mean ratios of incidence and reproduction numbers. We simulate 2000 epidemics under a reproduction number profile that has fixed mean of 2.5 up to *s* = 80 and then a step change fall to 0.5 until elimination, using the generation time distribution from [14]. We vary the offspring dispersion parameter *k* of the renewal model used to generate the epidemics and compute the variance-to-mean (VM) ratios of event reproduction numbers *R*_*s*_ and incidence *I*_*s*_ at every time *s*. In line with Eq. (4), there is a strong correspondence between VM[*R*_*s*_] and VM[*I*_*s*_] (mediated by the total infectiousness Λ_*s*_) and smaller *k* (larger heterogeneity) inflates both values.

This solidifies the idea of modulating VM[*I*_*s*_] via the control of VM[*R*_*s*_]. We consider three main strategies for implementing control in a heterogeneous epidemic. To make notation simpler we drop the subscript *s* and constrain all control protocols to reduce the mean reproduction number from *µ* to *ρµ* with *ρ* ≤ 1. Consequently, after control the new distribution describing *R* satisfies 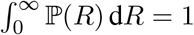 and 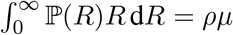 These descriptions equally model case reporting or ascertainment schemes, where *ρ* is the sample or reporting fraction instead [18].

i. Uniform control (constant reporting). Also called population-wide control and introduced in [18], this protocol assumes that all event reproduction numbers are reduced by the constant fraction *ρ*. This is achieved by rescaling Eq. (1) so that *R* ∼ Gam(*k, ρµk*^*−*1^) and is the simplest type of control that can be applied. Uniform control admits the analytic VM[*R*] = *ρµk*^*−*1^, which falls linearly as *ρ* → 0. It is equivalent to constant (or Bernoulli) reporting with the probability of observing or sampling any type of spreading event set at *ρ*.
ii. Super-spreading control (size-inverse reporting). Also called targeted control, and formulated in [7], this strategy disproportionately reduces super-spreading events and is within the class of individual-specific control measures detailed in [2]. It is implemented by upper-truncating the distribution of *R* at some maximum value *b* and rescaling so that 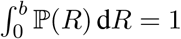 and 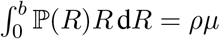. Here *b* defines the event *R* we consider associated with super-spreading. This is equivalent to size-inverse case ascertainment where super-spreading events are under- sampled. In this analogy *b* is the right under-reporting point i.e., we never observe events with *R > b*.
iii. Sub-spreading control (size-biased or preferential reporting). This a novel intervention that we introduce. It focuses on removing the sub-spreading events and is the converse of (ii). The gamma distribution of *R* is lower-truncated at some minimum value *a* and rescaled so that 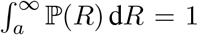 and 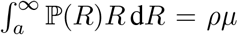. We use *a* to define sub-spreading events. This scheme is analogous to a size-biased case reporting strategy in which events producing few cases are under-sampled, with *a* as the left under-reporting point of the *R* distribution i.e., we never sample *R < a*. As (ii) and (iii) do not admit simple VM expressions we investigate them through simulation.

As *a* → 0 and *b* → ∞ all three control or reporting measures (and the scale parameter of their respective gamma *R* distributions) converge. They also converge as *k* → ∞ since the *R* distribution becomes degenerate at *ρµ* under these conditions and there is no heterogeneity. For simplicity, from this point we will usually refer to schemes (i)- (iii) via their control classification, switching to their reporting analogue only later when discussing results. We treat uniform control as a baseline since it ignores the specific form of the *R* distribution. While preferentially limiting super-spreading is sensible, and has been shown to have superior performance relative to uniform control [2], sub-spreading control has, to our knowledge, not been investigated. This is likely because it seems counter-intuitive to focus on events with low transmission potential.

However, we propose sub-spreading control based on the observation that a key source of the extra variability that over-dispersed distributions possess results from the increased probability of observing a zero sample i.e., zero inflation [32]. In an epidemic with heterogeneous transmission an excess of zero secondary cases would likely result from the sub-spreading event reproduction numbers. Here we investigate, for a fixed mean control effort *ρ*, whether the excess zeros or the extreme-tail events (i.e., the super-spreading ones) are more critical for shaping VM[*I*] (later we map VM[*I*] onto our elimination probabilities). We examine various *k* and compute the VM ratios for control strategies (i)–(iii) in Fig. 2 for increasing control effort i.e., decreasing *ρ*.

**Fig. 2:**
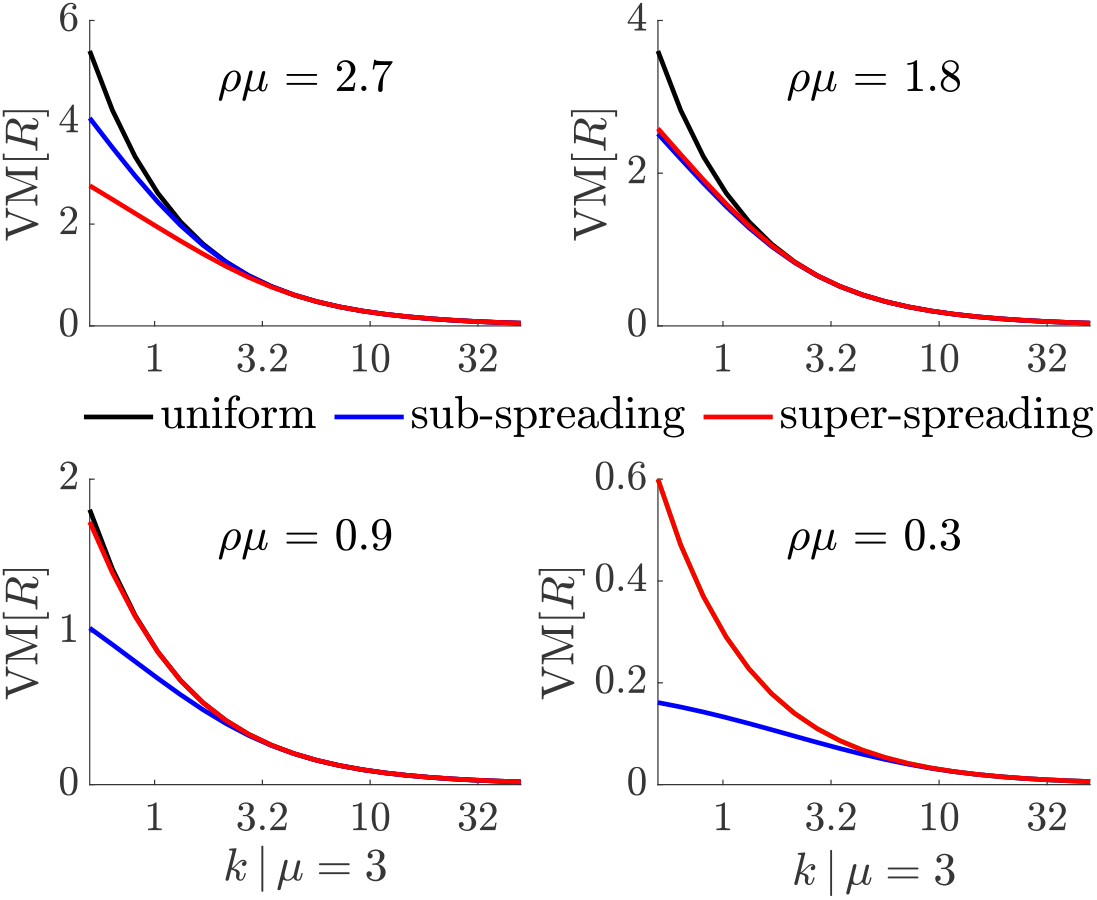
Control strategies and their variance-to-mean ratios. We compute the variance-to-mean (VM) ratios of the event reproduction number, *R*, distributions resulting from uniform, super-spreading and sub-spreading control measures for various dispersion parameters *k* with lower and upper truncations points of *a* = 1*/*10 and *b* = 10, respectively. Before control 𝔼[*R*] = *µ* = 3 and after control 𝔼[*R*] = *ρµ* with smaller *ρ* indicating increased average control effort. We find that when *ρµ* is large limiting super-spreading is most effective in reducing VM[*R*]. However, as *ρµ* decreases interventions targeting sub-spreading becomes the most effective.

Intriguingly, we find that when the controlled mean *R* i.e., *ρµ* is large, super-spreading control is most effective in reducing the VM ratio of the reproduction numbers. However, as the epidemic is better controlled and so *ρµ* falls, sub-spreading controls becomes the best intervention with respect to the VM ratio. This would make sense if as the mean controlled reproduction number falls the tail (super-spreading) events become increasingly improbable, meaning that most of the variability derives from the sub-spreading events. We confirm this notion in Fig. 3, which examines the cumulative distribution function *F* (*R*) at the extreme values of Fig. 2.

**Fig. 3:**
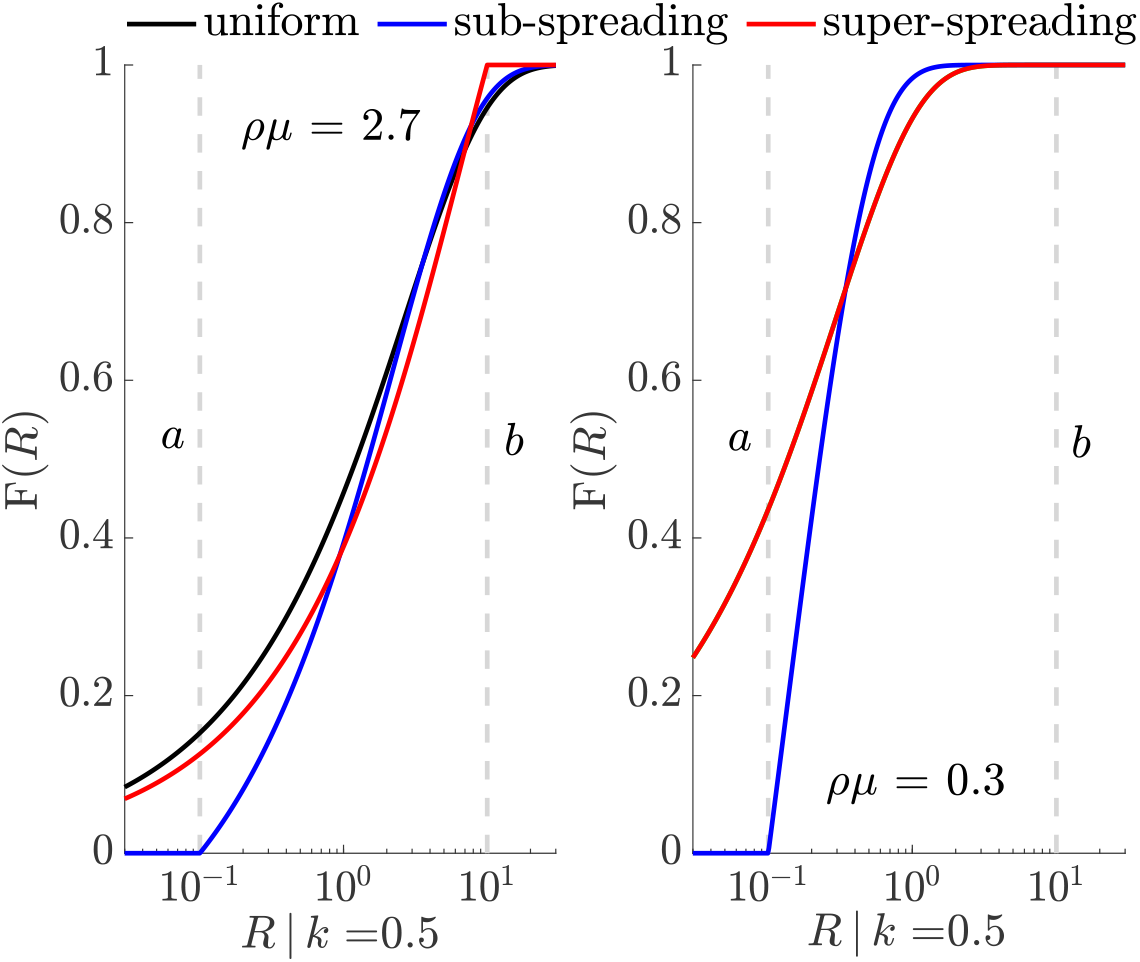
Control strategies shape reproduction number distributions. We plot the cumulative distribution function of the event reproduction number, *F* (*R*), for uniform, super-spreading (upper truncation *b*) and sub-spreading (lower truncation *a*) control measures under a renewal model with dispersion *k* = 0.5 at the largest and smallest mean controlled reproduction numbers, *ρµ*, examined in Fig. 2. We find that super-spreading control is best (in terms of variance-to-mean ratios) at large *ρµ* because there is a notable probability of super-spreading events that becomes truncated under this measure i.e., the resulting *F* (*R*) curve rises to 1 first due to *R > b* events being controlled. However, when *ρµ* is small this becomes vastly less important (the super-spreading and uniform controls converge) and the *F* (*R*) curve under sub-spreading control rises the fastest to 1 (where *R < a* events are suppressed).

Here we see that, at large *ρµ*, limiting super-spreading forces *F* (*R*) towards 1 at the fastest rate i.e., the clipping of super-spreading events closes the effective support of the *R* distribution earlier than the other measures. In this regime the tail events of the distribution are important. Elimination is not practically possible at such a large reproduction number, but this provides an interesting perspective for comparing to the low *ρµ* regime. There, because the tail events are strongly improbable, super-spreading and uniform control converge in behaviour. Interestingly, sub-spreading control forces *F* (*R*) more quickly to 1 than other measures, by truncating the small reproduction numbers. This may explain its performance in Fig. 2.

This assessment of the VM ratios and the effective reproduction number support (i.e., how quickly *F* (*R*) converges to 1) does not invalidate the results of [2], where a variant of super-spreading control increased VM ratios. We will resolve these apparent contradictions in a later section, highlighting the difference between renewal models and the branching processes used in [2]. We next investigate whether these reductions in VM[*R*] and VM[*I*] actually improve our ability to constrain *z*_*s*_ and the epidemic lifetime and hence to achieve reliable end-of-epidemic declaration times.

### Reducing variation among elimination times

We examine two data-justified measures of the end of an epidemic: the mean elimination time, 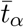 and the maximum elimination time *t*_*α*,max_. Both have previously been used for assessing end-of-epidemic declarations [13], [15]. As is convention, we focus on 95% confidence and set *α* = 95. Times 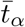 and *t*_*α*,max_ are obtained by finding when the mean and minimum of the *z*_*s*_ curves i.e., 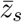 and *z*_*s*,min_, generated from possible future epidemic trajectories, first cross 0.95, respectively. We generate possible *z*_*s*_ by drawing samples from the distributions of 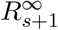 and then computing Eq. (6). All *z*_*s*_ curves incorporate the known past incidence 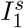 (see Methods).

We set 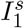 to the daily new infections observed over the MERS-CoV epidemic in South Korea in 2015, which was investigated in [29]. We then compute probabilities of zero case-days forward in time as in [15] but for various hypothetical *k* values and assess the elimination statistics using the 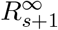 distributions of the control measures (i)–(iii) from above. In the appendix we perform complementary analyses using the incidence of the SARS 2003 epidemic in Hong Kong from [22] and simulated EVD data from [33], obtaining consistent results.

In Fig. 4 we present a range of mean and worst case *z*_*s*_ curves for various *k* with blue indicating the smallest *k* and red the largest. All times are given relative to the last observed non-zero case day. For all control measures we notice an interesting trend. First, and as expected from Eq. (8), we see that *k* ranks the mean *z*_*s*_ curves, with more heterogeneity (smaller *k*) leading to a larger 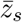. This behaviour is also consistent with [2], [30]. Second, we find that this ranking is inverted when we consider the worst case *z*_*s*,min_ instead i.e., epidemics with more heterogeneous transmission have larger worst case extinction or fade-out times.

**Fig. 4:**
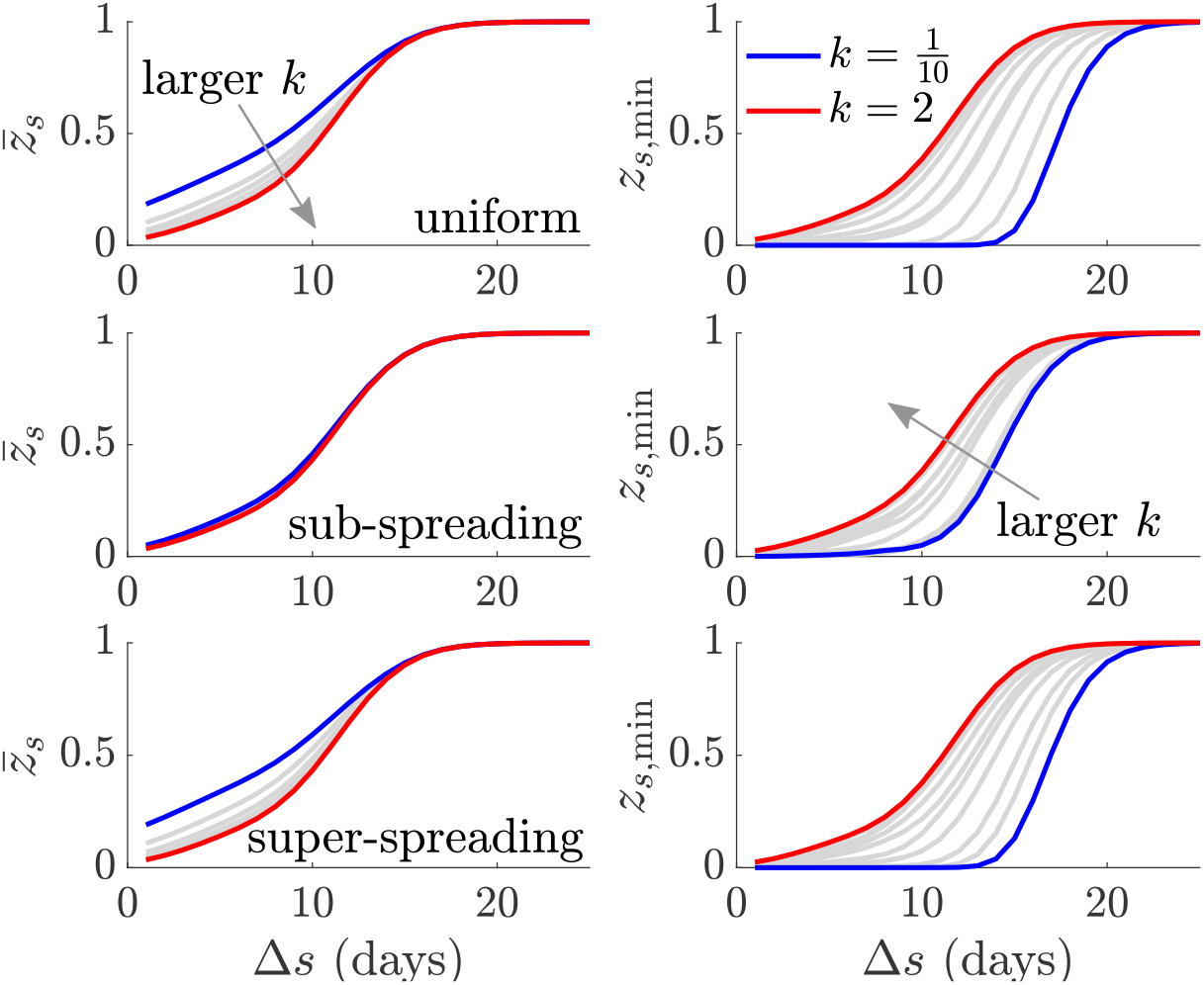
Mean and worst case epidemic elimination probabilities. We compute the mean 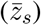 and worst case (*z*_*s*,min_) elimination probability curves using the incidence data from the MERS-CoV epidemic in South Korea in 2015 as in [29] i.e., we compute probabilities of sequences of zero future case days based on the available data and samples from the reproduction number distributions of the listed control scenarios (see main text). This procedure is repeated 2000 times to generate elimination curve distributions, from which we extract 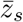 and *z*_*s*,min_. The curves are for various offspring dispersion parameters, *k*, rising from 0.1 to 2 under a mean controlled reproduction number of *ρµ* = 0.5. We find that increasing heterogeneity (smaller *k*) leads to earlier mean elimination times (the 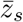 are larger) but later maximum (worst case) elimination times (the *z*_*s*,min_ are smaller).

This result is striking. It means that control actions which increase heterogeneity, and have been proposed as the most effective type of interventions [2], may lead to a false sense of confidence in the end of the epidemic. This follows from their larger worst case elimination times. Heterogeneity has the contrasting effect of contracting the mean lifetime of the epidemic but prolonging its maximum lifetime (counted in zero-case days). Since authorities would want a single decision time for safely declaring an epidemic to be over [16] and relaxing interventions (e.g., to re-open trade or travel), this creates a practical, potentially costly and unexpected complication.

However, we observe that non-uniform control measures can help alleviate this problem. The sub-spreading control scheme considerably shrinks the variation among *z*_*s*_ curves. This proceeds from the results of the previous section, where we found that, at small mean reproduction numbers (which is realistic when we are near the tail or end of an epidemic), limiting sub-spreading significantly reduces VM[*R*_*s*_] and hence VM[*I*_*s*_] (see Eq. (4)). We assess this in more detail by examining all the *z*_*s*_ curves, which led to the means and minima in Fig. 4, and the distributions of the 95% declaration times, *t*_95_, that result. We provide these in Fig. 5.

**Fig. 5:**
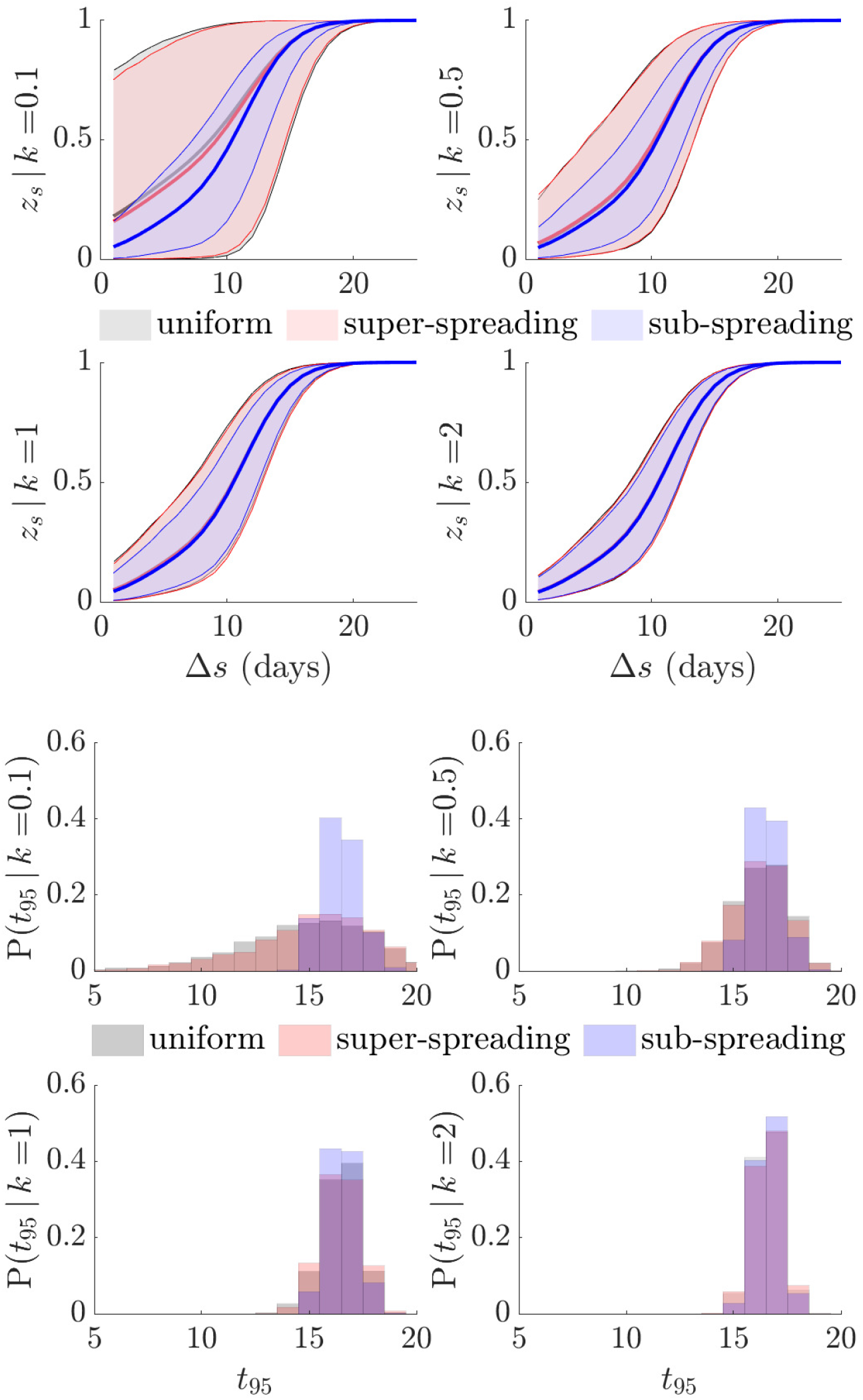
Elimination curves and declaration times for various control strategies. We compute elimination probabilities (*z*_*s*_) of future trajectories of zero case-days starting from the incidence data of the MERS-CoV epidemic in South Korea in 2015 and in line with [29]. Each trajectory is formed by sampling from the future reproduction number, 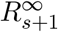, distributions for uniform, super-spreading and sub-spreading control measures. This procedure is repeated for 2000 possible trajectories. The top panel shows the *z*_*s*_ distribution from these trajectories for various offspring dispersion parameters *k* and the bottom panel provides corresponding 95% declaration times (*t*_95_). All times are relative to that of the last observed case and we use a mean controlled reproduction number of *ρµ* = 0.5 at every future time. We find that sub-spreading control is most effective at reducing the variability (or equivalently increasing the reliability) of both *z*_*s*_ and *t*_95_. We bolster this assertion with additional examples in Fig. A.1.

As *k* becomes larger the difference among all control measures expectedly shrinks. For epidemics with significant heterogeneity (*k <* 1) we observe that both uniform and super-spreading control result in large variations in *z*_*s*_ (top red and grey curves in Fig. 5, which mostly overlay each other). This manifests in a notable spread of 95% declaration times (bottom red and grey bars of Fig. 5, also overlaid). Sub-spreading control is, however, able to suppress much of this variation yielding more deterministic elimination probabilities and declaration times (blue curves and bars in Fig. 5) and thus minimising the possibility of early or late declarations.

We observe consistency in this trend for both EVD and SARS incidence curves in Fig. A.1 of the appendix. We further confirm the ability of sub-spreading control as a means of making heterogeneous incidence curves more deterministic in behaviour by simulating complete epidemic trajectories under each control measure in the bottom panel of Fig. 6. There we see that the VM ratios of the incidence are indeed minimised by sub-spreading control measures (the mean incidence from all control measures there is approximately the same).

**Fig. 6:**
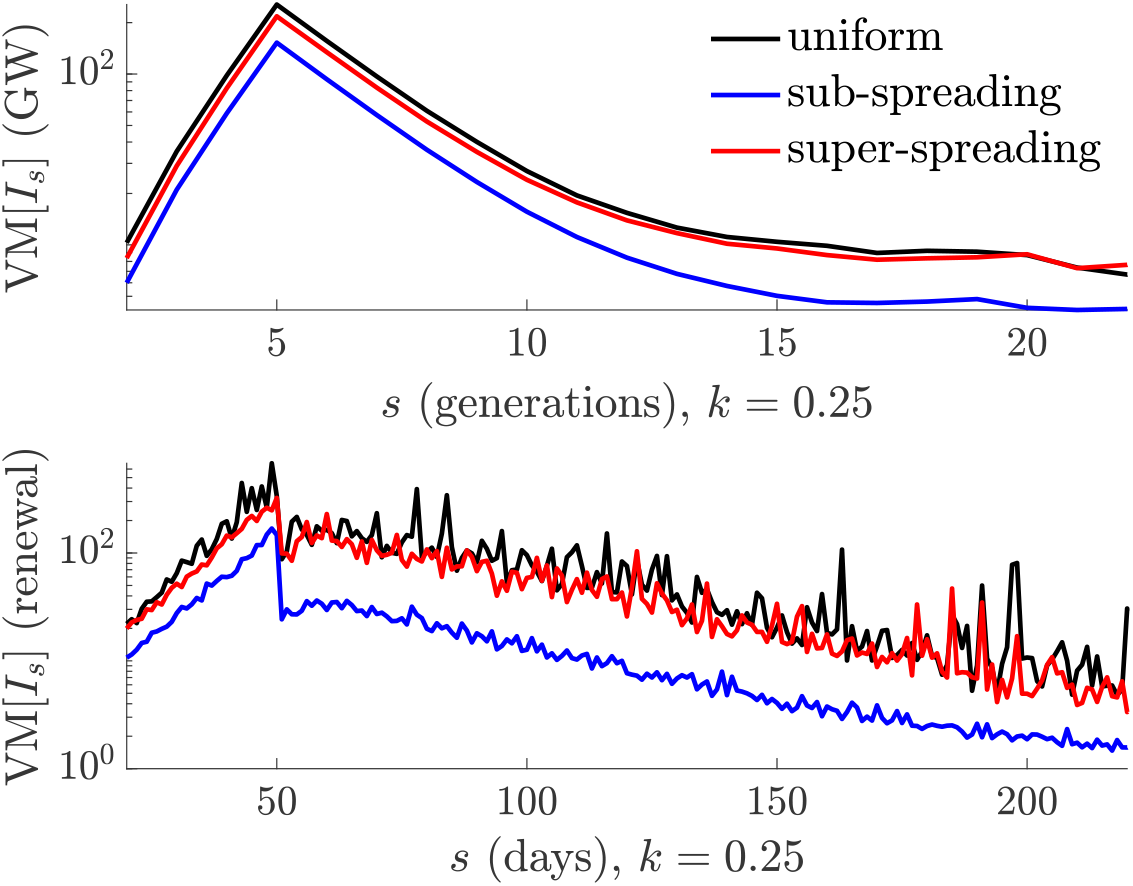
VM ratios for simulated epidemics. Using either a Galton-Watson (GW) branching process [2] (top) or a renewal model [21] (bottom) we simulate 10^4^ trajectories under a step-change in mean effective reproduction number, *ρµ*, from 2.5 to 0.6, with dispersion *k* = 0.25. At each time point we draw event reproduction numbers from the uniform, super-spreading and sub-spreading distributions with these means. We consistently find, for waning epidemics, that sub-spreading minimises variance-to-mean (VM) ratios of the incidence (denoted *I*_*s*_ at time *s*). The mean incidence from all methods is approximately the same.

At this point, we re-interpret these results from the perspective of case ascertainment. The similarity of super-spreading and uniform control mean that reporting strategies that sample possible spreading events with constant probability or inversely to their reproduction numbers have approximately the same effect on elimination waiting times. This likely follows from the vanishingly low probability of super-spreading. However, size-biased sampling, which is not only the analogue to sub-spreading control, but also likely to occur in practice [18], has an important effect. Failing to observe sub-spreading events leads to a strongly overconfident view of elimination. The epidemic tail appears far less variable when those events are excluded, leading to risky end-of-epidemic declarations.

Last, we comment on differences between our renewal model approach and the Galton-Watson (GW) branching process used in [2]. Both models have significant dynamical disparities. Specifically, GW processes are truly only valid early in an epidemic, use a deterministic generation time and assume that the number of infected in the next generation only depend on those in the current generation. These characteristics make the GW process unsuitable for the analysis of elimination statistics [7], especially when the generation time distribution is known (a) to be non-degenerate, (b) to have long memory and (c) to determine the epidemic growth rate [12], [20], [34].

Further, our problem of interest is ill-defined for GW processes since, under these models, we always obtain *t*_95_ = 1 (time is now in generations). This follows as 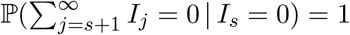. Our results therefore do not apply to GW processes and should not be expected to correspond with computations of the probability of extinction from [2]. Nevertheless, we make some key comparisons. In [2], increasing heterogeneity elevated the extinction probability of an epidemic by promoting early burn-out. This partially holds true for renewal models, but (a) mostly affects *t*_0_ and (b) is not meaningful here as we condition on a significant epidemic having existed.

Since elimination measures only become an important consideration after *t*_0_ and for epidemics of notable size [16], [29], burn-out due to overdispersion does not benefit our analysis. The extra dynamics beyond *t*_0_, which do not exist for GW processes, form our problem of interest. However, the VM ratio behaviour of our control schemes (i)–(iii) is quite general and still works under GW models. In Fig. 6 we show that the GW process (top panel) conserves the ordering of VM ratios from the renewal model (bottom panel), hence confirming the relative importance of sub-spreading control for achieving reliable elimination.

## Discussion

Understanding how transmission heterogeneity shapes the dynamics of diverse phases of an emerging epidemic is crucial if we are to safely and efficiently apply or relax interventions during those phases. While much research has focussed on super-spreading and the growing phases of epidemics, sub-spreading and the dynamics of waning infectious diseases have been understudied [9], [30]. In this paper we investigated both, by developing a general framework to measure how the variation or heterogeneity (embodied by *k*) around mean transmission at some time *s* (summarised by *µ*_*s*_, the mean of *R*_*s*_), engenders risk in the probability of epidemic elimination, *z*_*s*_.

We extended the popular theory of transmission heterogeneity from [2] to precisely describe how variations (via VM ratios) in event reproduction numbers, *R*_*s*_, manifest in fluctuations of the incidence, *I*_*s*_ (see Fig. 1). The resulting proportional relationship meant that the *R*_*s*_ distribution directly modulates the observed *I*_*s*_ curve [24]. We then generalised the formulae of [3], [15] to derive how these variations propagate uncertainty onto *z*_*s*_. Since end-of-epidemic declarations depend on when *z*_*s*_ crosses a confidence threshold, this uncertainty makes adjudging when an epidemic is over complex, risky and potentially costly [13], [16].

Using our framework, we found that when epidemics are strongly heterogeneous, i.e., *k <* 1, the maximum and mean declaration times, relative to that of the last observed case, depend contrastingly on *k* (see Fig. 4). Although the average time to declaration decreases with *k*, supporting previous work linking heterogeneity to epidemic extinction [14], the concomitant increase in variability means that the safest declaration time actually increases and the risk of early or late declarations can be severely amplified. Variation originating from transmission heterogeneity is therefore not beneficial for achieving safe and reliable end-of-epidemic declarations.

Consequently, we investigated if targeted control can ameliorate this end-of-epidemic volatility. We considered three control schemes (for the same mean level of control *ρ*), which were non-selective or targeted either super-or sub-spreading (see Fig. 2 and Fig. 3). Intriguingly, we found that, because the controlled mean of the event reproduction numbers *ρµ*_*s*_ is below 1 at pre-elimination settings, curbing super-spreading only marginally improved on non-selective approaches. However, sub-spreading appeared to be the main contributor to end-of-epidemic declaration risk, meaning limiting those events can significantly reduce that risk; forcing the epidemic tail to be more deterministic and increasing the reliability of resulting intervention relaxation policies.

This result, while new and surprising, is supported by current understanding. Overdispersed distributions have two defining characteristics: tall heads and fat tails [24], where head and tail refer to the left and right ends of our *R*_*s*_ distributions. Since *ρµ*_*s*_ *<* 1 near elimination and because we only start measuring time once a sequence of days with no cases appear (in accordance with official guidelines [16]) fat tail events are extremely unlikely to affect epidemic dynamics. Consequently, it is the tall head i.e., the excess of events with *R*_*s*_ ≪ *ρµ*_*s*_, which are responsible for sustained overdispersion. Curbing sub-spreading suppresses this remaining source of risk.

We validated these theories on empirical MERS-CoV, SARS and simulated EVD epidemics (where we computed forward probabilities of no future cases based on these given data) and observed significant reductions in end-of-epidemic declaration time spread (see Fig. 5 and Fig. A.1) when sub-spreading was controlled. The conditions under which these effects were prominent (when *k <* 0.5) are realistic for these and many other infectious diseases [2], [26]. An efficient strategy for controlling and then reliably eliminating an epidemic might therefore re-direct interventions from targeting super-spreading to sub-spreading transmission as infections enter waning phases.

Unfortunately, such selective strategies may be infeasible or difficult to realise. Predicting or simply identifying specific spreading event-types has proven difficult and hampered attempts at targeting super-spreading [8]. As sub-spreading frequently involves zero secondary cases, these events may be significantly harder to investigate. Intensive contact tracing and isolation schemes can help ameliorate some of these issues. Ongoing studies into associations among measurable traits, for example viral loads or symptom severity, and infectiousness or transmissibility, may also make identifying and curbing specific spreading events easier [35], [36]. However, even if these identification problems are overcome, policy resistance can nullify the benefits of targeted strategies [37].

Despite these points, our analysis of spreading events and our framework have several practical implications. First, we showed that the benefits of curbing super-spreading diminish with incidence. If targeted control is more costly than population-wide surveillance measures, this can support policy switches as the epidemic wanes. Second, we exposed the inherent risk of control measures that neglect sub-spreading. This knowledge can evidence more risk-averse approaches to end-of-epidemic declarations, in line with works such as [14]. Third, key formulae within our framework (Eq. (2) – Eq. (8) and Eq. (10)) are valid for arbitrary reproduction number distributions. Consequently, we can test the influence of other types of heterogeneity (e.g., demographic or spatial effects) on epidemic lifetimes and even incorporate empirical effective reproduction number distributions when available.

An important consequence of our analyses emerges from the mathematical analogue between control and case ascertainment [18]. Uniform, super-spreading and sub-spreading control are equivalent to constant, size-inverse and size-biased reporting. Size-biased reporting protocols are common in practice and generally include any preferential sampling scheme that observes infectious events with likelihoods that grow with the number of secondary cases produced by that event [24], [38]. We investigated a specific size-biased scheme, similar to that in [18], where sub-spreading events are unobserved. This leads (by analogy to our sub-spreading control results) to a drastic underestimation of the variance of possible elimination times exhibited by heterogeneous epidemics.

Previously it was shown that the mean end-of-epidemic declaration time is biased by any type of under-reporting [15]. As a result, practical surveillance schemes, which will almost surely feature some degree of size-biased reporting, are likely to promote simultaneously biased and overconfident end-of-epidemic declarations. These effects emphasise why sustained high quality epidemic monitoring is crucial for guiding the relaxation of interventions or the reopening of economies during the waning phases of outbreaks. In the absence of such monitoring our results recommend a more cautious approach to designating the end of an outbreak.

While the results we have presented expose the complexities and biases in adjudging epidemic fade-out, there are some limitations to our analyses. We have assumed that the control effort or reporting rate of *ρ* can be realised with equal ease for both the uniform and selective strategies and that gamma or truncated gamma reproduction number distributions are sensible. These are necessary and standard assumptions for comparing strategies and sensibly summarising heterogeneity [2], [7] but their validity may depend on the disease being considered and the properties (e.g., contact networks) of the area under study. Scale may also matter. For example, across larger regions population-wide measures may more easily achieve a given *ρ* than more targeted ones.

Moreover, we have not accounted for how interventions (e.g., lockdowns or quarantines) may alter the contact networks and behaviours of individuals and hence the characteristics of serial interval or generation time and offspring distributions (which may vary across epidemic phases) [39]. Such changes could affect the adequacy of the renewal models we have employed or the practical effectiveness of the strategies we have examined. While lack of data precludes improvement here, we note that our framework for investigating epidemic fade-out (a) is sufficiently general to handle empirical (and non-gamma) reproduction number distributions if such knowledge is available, (b) remains valid if up-to-date serial intervals and dispersion parameters are used and (c) applies to the waning phases of the epidemic, after most drastic changes would likely have already settled.

Our framework provides a general toolkit for testing hypotheses about targeted controls or case ascertainment schemes and measuring their influence on end-of-epidemic declaration times. It can also be easily extended to include additional factors such as imported cases or to investigate other types of heterogeneity (e.g., age-based reproduction numbers) [15]. As the current COVID-19 pandemic underscores, much still remains unknown about the relative merits of elimination approaches, e.g., “zero COVID” strategies [11], and the influence of heterogeneity [4], [40]. Improved understanding of epidemic dynamics can only aid preparedness and decision-making. We hope that our framework, which exposed unexpected consequences of understudied spreading events [19], can contribute towards this goal and help inform safe intervention relaxation and end-of-epidemic declaration strategies.

## Data Availability

All data and code are freely available at https://github.com/kpzoo/sub-spreading

## Funding

This work is jointly funded under grant reference MR/R015600/1 by the UK Medical Research Council (MRC) and the UK Department for International Development (DFID) under the MRC/DFID Concordat agreement and is also part of the EDCTP2 programme supported by the European Union.

## Appendix

### A. Declaration time histograms for SARS and EVD

**Fig. A.1:**
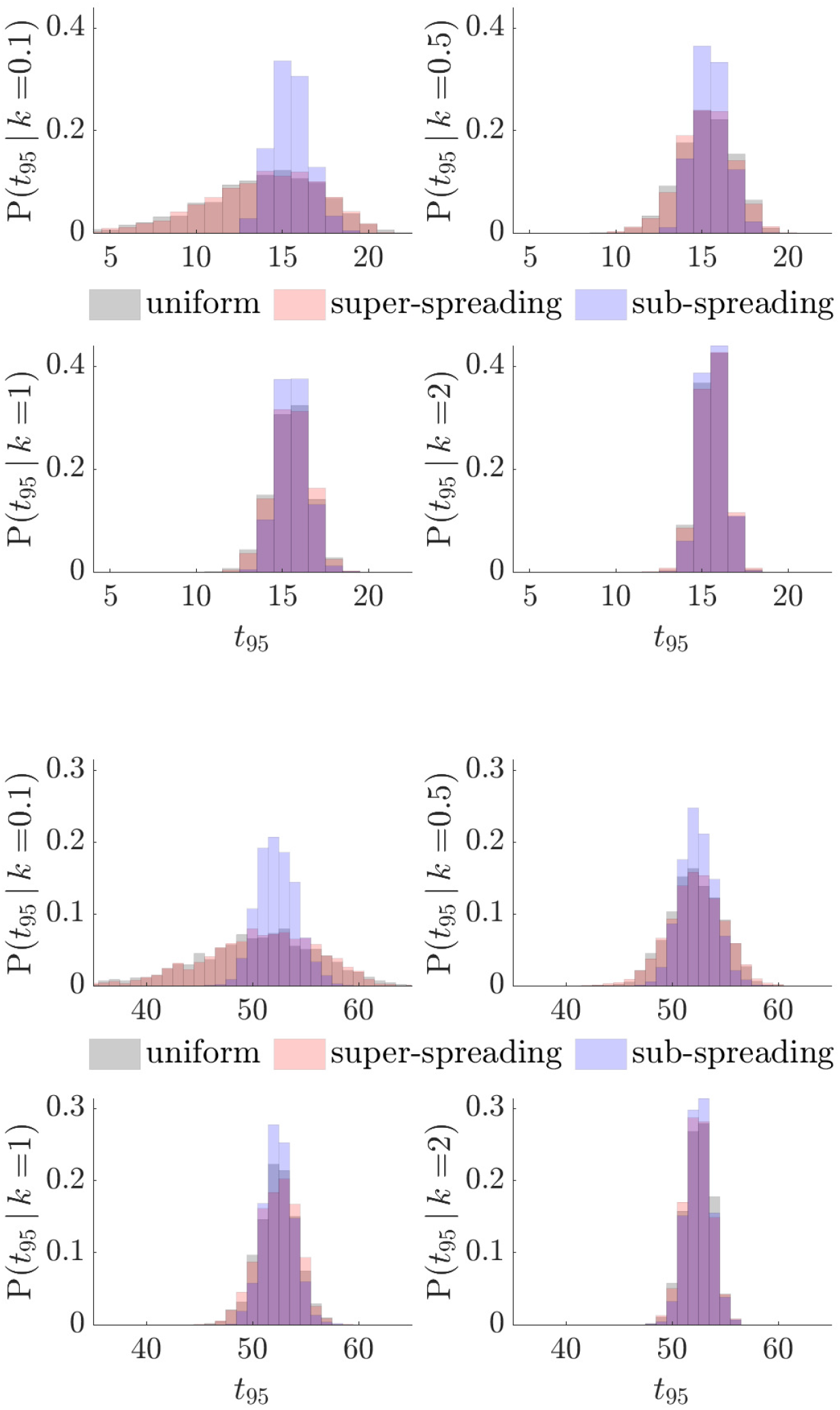
Declaration times for different epidemics. We generate 2000 future trajectories of zero case-days using empirical incidence data of the SARS epidemic in Hong Kong in 2003 (top) [22] and simulated incidence for EVD (bottom) from [33]. We obtain trajectories by sampling from the future reproduction number, 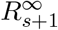, distributions for uniform, super-spreading and sub-spreading control measures and plot corresponding 95% declaration times (*t*_95_). The uniform and super-spreading results largely overlap. All times are relative to that of the last observed case and assume a mean controlled reproduction number of *ρµ* = 0.5. As in the main text, we see that sub-spreading control is most effective at reducing the variability (increasing the reliability) of end-of-epidemic declarations.

## Notes

### Competing Interest Statement

The authors have declared no competing interest.

### Summary of Updates

Improved figures and reconfigured text.

## References

[1] Nishiura H, Chowell G. The Effective Reproduction Number as a Prelude to Statistical Estimation of Time-Dependent Epidemic Trends. In: Mathematical and statistical estimation approaches in epidemiology. Springer; 2009. p. 103–21.

[2] Lloyd-Smith J, Schreiber S, Kopp P, et al. Superspreading and the effect of individual variation on disease emergence. Nature. 2005;438(17):355–9.

[3] Parag K, Cowling B, Donnelly C. Deciphering early-warning signals of the elimination and resurgence potential of SARS-CoV-2 from limited data at multiple scales. medRxiv. 2021;2020.11.23.20236968.

[4] Susswein Z, Bansal S. Characterizing superspreading of SARS-CoV-2 : from mechanism to measurement. medRxiv. 2020;2020.12.08.20246082.

[5] De Serres G, Gay N, Farrington P. Epidemiology of Transmissible Diseases after Elimination. Am J Epidemiol. 2000;151(11).

[6] Woolhouse M, Dye C, Etard J, et al. Heterogeneities in the transmission of infectious agents: Implications for the design of control programs. PNAS. 1997;94:338–42.

[7] Garske T, Rhodes C. The effect of superspreading on epidemic outbreak size distributions. J Theor Biol. 2008;253:228–37.

[8] Frieden T, Lee C. Identifying and Interrupting Superspreading Events— Implications for Control of Severe Acute Respiratory Syndrome Coronavirus 2. Emerg Infect Dis. 2020;26(6):1059–66.

[9] Britton T, House T, Loyd A, et al. Five challenges for stochastic epidemic models involving global transmission. Epidemics. 2015;10:54– 7.

[10] Parag K. Improved estimation of time-varying reproduction numbers at low case incidence and between epidemic waves. medRxiv. 2020;2020.09.14.20194589.

[11] Baker M, Wilson N, Blakely T. Elimination may be the optimal response strategy for COVID-19 and other emerging pandemic diseases. BMJ. 2020;371:m4907.

[12] Lee H, Nishiura H. Sexual transmission and the probability of an end of the Ebola virus disease epidemic. J Theor Biol. 2019;471:1–12.

[13] Nishiura H, Miyamatsu Y, Mizumoto K. Objective determination of end of MERS outbreak, South Korea. Emerg Infect Dis. 2016;22:146–8.

[14] Djaafara B, Imai N, Hamblion E, et al. A quantitative framework to define the end of an outbreak: application to Ebola Virus Disease. Am J Epidemiol. 2020;p. kwaa212.

[15] Parag K, Donnelly C, Jha R, et al. An exact method for quantifying the reliability of end-of-epidemic declarations in real time. PLOS Comput Biol. 2020;16(11):e1008478.

[16] WHO. WHO recommended criteria for declaring the end of the Ebola virus disease outbreak; 2020. Available from: https://www.who.int/who-documents-detail/who-recommended-criteria-for-declaring-the-end-of-the-ebola-virus-disease-outbreak.

[17] Lloyd-Smith J, Cross P, Briggs C, et al. Should we expect population thresholds for wildlife disease? Trends Ecol Evol. 2005;20(9):511– 19.

[18] Lloyd-Smith J. Maximum Likelihood Estimation of the Negative Binomial Dispersion Parameter for Highly Overdispersed Data, with Applications to Infectious Diseases. PLOS One. 2007;2:e180.

[19] Lawyer G. Understanding the influence of all nodes in a network. Sci Rep. 2015;5(8665).

[20] Wallinga J, Lipsitch M. How generation intervals shape the relationship between growth rates and reproductive numbers. Proc R Soc B. 2007;274:599–604.

[21] Fraser C, Cummings D, Klinkenberg D, et al. Influenza Transmission in Households During the 1918 Pandemic. Am J Epidemiol. 2011;174(5):505–14.

[22] Cori A, Ferguson N, Fraser C, et al. A New Framework and Software to Estimate Time-Varying Reproduction Numbers During Epidemics. Am J Epidemiol. 2013;178(9):1505–12.

[23] Parag K, Donnelly C. Adaptive Estimation for Epidemic Renewal and Phylogenetic Skyline Models. Syst Biol. 2020;69(6):1163–79.

[24] Karlis D, Xekalaki E. Mixed Poisson Distributions. Intern Statist Rev. 2005;73(1):35–58.

[25] Churcher T, Cohen J, Ntshalintshali N, et al. Measuring the path toward malaria elimination. Science. 2014;344(6189):1230–32.

[26] Lau M, Dalziel B, Funk S, et al. Spatial and temporal dynamics of superspreading events in the 2014-2015 West Africa Ebola epidemic. PNAS. 2017;114(9):2337–42.

[27] Parag K, Donnelly C. Using information theory to optimise epidemic models for real-time prediction and estimation. PLOS Comput Biol. 2020;16(7):e1007990.

[28] Azmon A, Faes C, Hens N. On the estimation of the reproduction number based on misreported epidemic data. Stats Med. 2014;33:1176– 92.

[29] Nishiura H. Methods to determine the end of an infectious disease epidemic: a short review. In: Chowell G, Hyman J, editors. Mathematical and statistical modeling for emerging and re-emerging infectious diseases; 2016. p. 291–301.

[30] Yan P, Chowell G. Quantitative Methods for Investigating Infectious Disease Outbreaks. vol. 70 of Texts in Applied Mathematics. Cham, Switzerland: Springer; 2019.

[31] Wasserman L. All of Statistics: A Concise Course in Statistical Inference,. Berlin, Germany: Springer-Verlag; 2003.

[32] Xekalaki E. On the distribution theory of over-dispersion. J Statistic Distrib Applic. 2014;1(19).

[33] Jombart T, Frost S, Nouvellet P, et al. outbreaks: A Collection of Disease Outbreak Data; 2019 [cited 31 Jul 2020]. Available from: https://github.com/reconhub/outbreaks.

[34] Parag K, Thompson R, Donnelly C. Are epidemic growth rates more informative than reproduction numbers? medRxiv. 2021;2021.04.15.21255565.

[35] Edwards A, Ausiello D, Salzman J, et al. Exhaled aerosol increases with COVID-19 infection, age, and obesity. PNAS. 2021;118(8):e2021830118.

[36] Goyal A, Reeves D, Cardozo-Ojeda E, et al. Viral load and contact heterogeneity predict SARS-CoV-2 transmission and super-spreading events. eLife. 2021;10:e63537.

[37] Wang A, Bauch C, Bhattacharyya S, et al. Statistical physics of vaccination. Phys Rep. 2016;664:1–113.

[38] Zelner J, Masters N, Broen K, et al. Preferential observation of large infectious disease outbreaks leads to consistent overestimation of intervention efficacy. medRxiv. 2020;2020.11.02.20224832.

[39] Ali S, Wang L, Lau E, et al. Serial interval of SARS-CoV-2 was shortened over time by nonpharmaceutical interventions. Science. 2020;369(6507):1106–9.

[40] Lewis D. Superspreading drives the COVID pandemic — and could help to tame it. Nature. 2021;590:544–6.

